# ChatGPT-CARE: a Superior Decision Support Tool Enhancing ChatGPT with Clinical Practice Guidelines

**DOI:** 10.1101/2023.08.09.23293890

**Authors:** Yanshan Wang, Shyam Visweswaran, Sumit Kapoor, Shravan Kooragayalu, Xizhi Wu

## Abstract

ChatGPT has gained remarkable traction since its inception in November 2022. However, it faces limitations in generating inaccurate responses, ignoring existing guidelines, and lacking reasoning when applied in clinical settings. This study introduces ChatGPT-CARE, a tool that integrates clinical practice guidelines with ChatGPT, focusing on COVID-19 outpatient treatment decisions. By employing in-context learning and chain-of-thought prompting techniques, ChatGPT-CARE enhances original ChatGPT’s clinical decision support and reasoning capabilities. We created three categories of various descriptions of patients seeking COVID-19 treatment to evaluate the proposed tool, and asked two physicians specialized in pulmonary disease and critical care to assess the responses for accuracy, hallucination, and clarity. The results indicate that ChatGPT-CARE offers increased accuracy and clarity, with moderate hallucination, compared to the original ChatGPT. The proposal ChatGPT-CARE could be a viable AI-driven clinical decision support tool superior to ChatGPT, with potential applications beyond COVID-19 treatment decision support.

## Introduction

ChatGPT has witnessed impressive growth since its initial launch in November 2022, amassing an estimated monthly user base of more than 100 million^1^. ChatGPT is an artificial intelligence (AI) technology that is grounded in a comprehensive unsupervised learning process that utilizes massive amounts of text data from the internet. At the time of writing, there are two versions of ChatGPT in use: GPT-3.5 and GPT-4. The former, GPT-3.5, is an improvement over the previous version, GPT-3, and incorporates reinforcement learning strategies to improve its capabilities as a conversational agent, particularly in terms of providing accurate and helpful responses to users’ queries. The specific foundational characteristics of GPT-4 have not been made public. However, according to recent literature^2^, GPT-4 is thought to be a substantial advance over GPT-3.5. This advance is primarily measured by its increased accuracy and diminished propensity for generating responses that are unrelated to the input and sometimes contain misinformation, a phenomenon known in the field of generative AI as “hallucination.” GPT-3.5 and GPT-4 are both based on Generative Pretrained Transformer (GPT) models. Since they have billions of parameters within the GPT neural network architectures, they are known as large language models (LLMs).

Due to its impressive capabilities in generating coherent responses to input queries, ChatGPT has shown its potential to revolutionize clinical care and clinical workflow^3^. It has already been extensively evaluated in a variety of settings to assist healthcare providers. Among these applications are those that aid clinical decision making^4–6^, automate clinical documentation^7^, expedite prior authorization^3^, and enhance patient communication^8^. Despite its great promise, ChatGPT has significant limitations. It can sometimes ignore important contextual information^9^, generating inaccurate responses^10^. Moreover, the generated response may be overly general, lack transparency in decision-making reasoning, and may not align with clinical practice guidelines. These constraints currently limit its widespread use in clinical applications.

In this study, we integrated clinical practice guidelines with ChatGPT and built a new tool called ChatGPT-CARE. Through a thorough review by two physicians, we demonstrate that this new tool could enhance the capabilities of ChatGPT for clinician decision support. We specifically focused on evaluating ChatGPT-CARE’s performance in supporting outpatient treatment decisions for Coronavirus Disease 2019 (COVID-19). Because COVID-19 was a novel disease at the start of the pandemic, clinical practice guidelines evolved rapidly in response to new discoveries and treatments. Keeping up with the frequent guideline updates was difficult for healthcare providers. In such a situation, a clinical decision support tool like ChatGPT-CARE can be valuable in providing reliable support for decision making in clinical care.

## Methods

### COVID-19 Outpatient Treatment Guidelines

For the COVID-19 outpatient treatment guidelines, we used the Centers for Disease Control and Prevention (CDC) and Infectious Diseases Society of America (IDSA) COVID-19 Outpatient Treatment Guidelines, version 10 (updated on February 2, 2023)^11^. The Guidelines provide a comprehensive step-by-step approach that outlines outpatient treatment options. We revised the Guidelines slightly to reflect insights and recommendations from our physicians. Body weight, in particular, was used as a key determinant for COVID-19 treatment since many medication dosages are weight-based. We considered the patient’s underlying medical conditions, COVID-19 related symptoms and the duration of symptoms to determine if they were in the high-risk versus low-risk category. Based on this information, the Guidelines offer four treatment options for low-risk patients: Paxlovid, Remdesivir, Molnupiravir, or supportive care. The revised COVID-19 Outpatient Treatment Guidelines were used to build ChatGPT-CARE for COVID-19 outpatient treatment decision support and can be found in the Supplementary Appendix.

### In-context Learning

ChatGPT-CARE utilizes in-context learning to incorporate clinical practice guidelines and enhance ChatGPT’s decision support capabilities. Since most LLMs have billions of parameters, additional model training necessitates extensive computation and, in most cases, is impractical. Through the incorporation of pre-defined prompts, in-context learning has emerged as a new paradigm for fine-tuning LLMs without additional training for specific tasks. Typically, the pre-defined prompts begin with a system prompt, followed by a few alternating input and output prompts (also known as few-shot learning). The system prompt establishes ChatGPT’s role or function, whereas the few-shot learning prompts provide examples of desired output. Therefore, in ChatGPT-CARE, the process of in-context learning consists of two steps: 1) a system prompt that integrates the revised COVID-19 Outpatient Treatment Guidelines into ChatGPT; and 2) several few-shot learning prompts that fine-tune the model for COVID-19 treatment decision support. Following this approach, ChatGPT-CARE becomes a powerful tool for providing accurate COVID-19 treatment suggestions along with a transparent decision making process. Figure 1 depicts the ChatGPT-CARE tool’s in-context learning process and treatment decision support.

**Figure 1.**
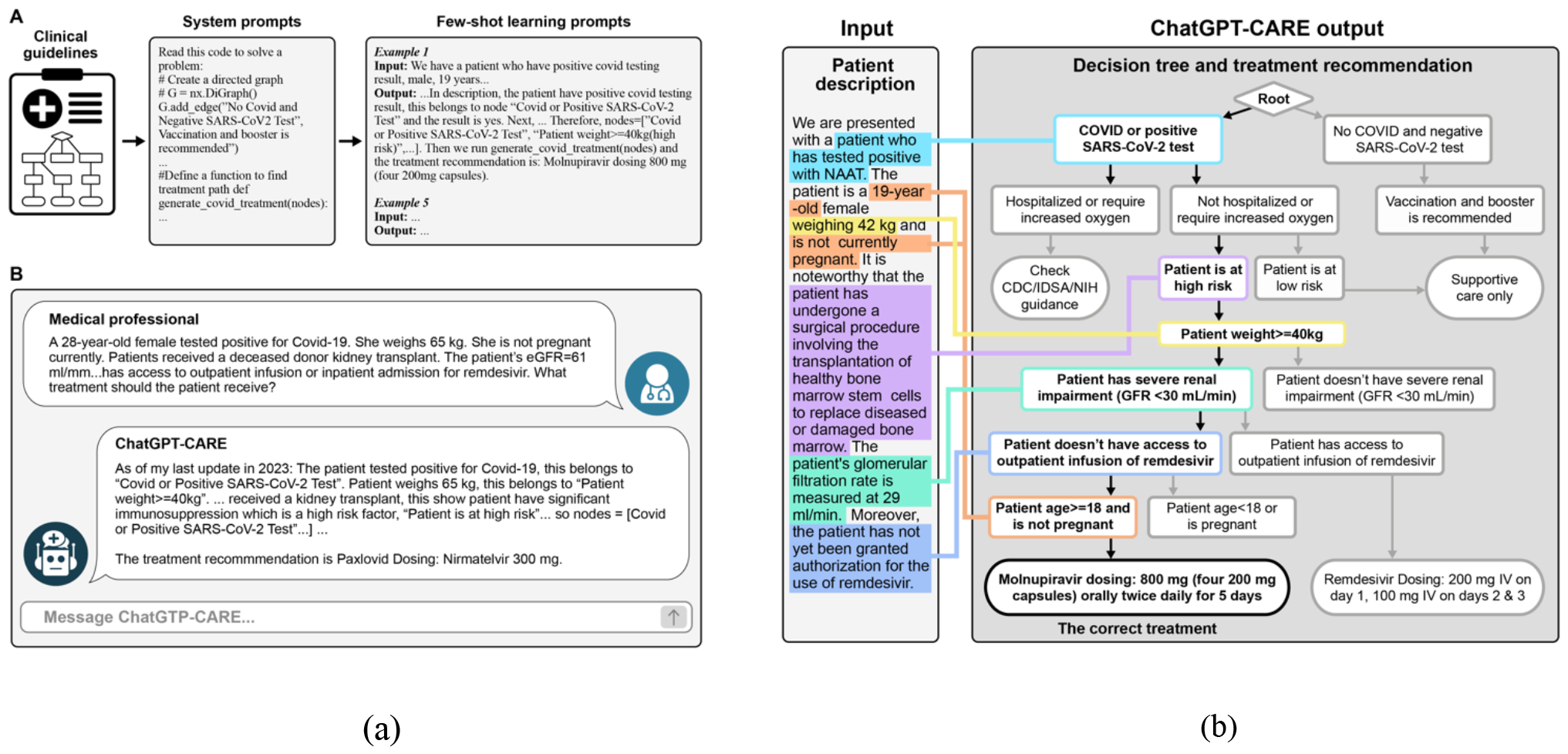
Illustration of ChatGPT-CARE for COVID-19 outpatient treatment decision support. (a) shows the process of knowledge engineering of clinical practice guidelines into ChatGPT. (b) shows ChatGPT’s decision making process given an example of patient description.

#### System Prompt: Clinical Practice Guideline as Coded Graph

In order to incorporate the knowledge of COVID-19 Outpatient Treatment Guidelines into ChatGPT, we transformed the guidelines into a Python-coded graph prompt. In the prompt, we used the NetworkX^12^ library to construct the Guidelines as a directed knowledge graph, with nodes representing medical condition checkpoints or final treatment options and edges representing possible transitions between these nodes. In addition, the system prompt consists of a Python function that is responsible for determining the path to the treatment options based on the given nodes as input. ChatGPT-CARE uses this function to find a path to a treatment option and return the COVID-19 patient’s recommended

#### Few-shots Learning Prompt: Chain-of-thought Prompting for Reasoning

We adopted the few-shots learning strategy to refine ChatGPT-CARE. Few-shots learning is typically used to fine-tune LLMs with a small number of annotated data points. It is useful when obtaining extensive annotated data becomes difficult, time-consuming, or impractical, particularly in real-world healthcare-related Natural Language Processing (NLP) tasks^13^. In ChatGPT-CARE, We used 5-shot learning, which means that the model was fine-tuned with five input and output examples. Each example started with a patient description followed by the chain-of-thought prompting response. The rationale behind selecting five examples was to encompass distinct situations, ensuring comprehensive learning across various scenarios. The first example used the longest path in the Python-coded graph, which we consider to be the most difficult case for ChatGPT-CARE among all possible treatment recommendations. The second and third examples are shorter paths with only three and four nodes, respectively. We discovered that this could teach ChatGPT-CARE how to handle shorter paths more effectively. The fourth and fifth examples were derived from the errors encountered during the earlier development stages of ChatGPT-CARE. We discovered that including these erroneous examples in our prompts could help the model avoid making similar mistakes in the future. Furthermore, we deliberately injected unrelated and subtle information into all five questions, providing instructions in the responses and explanations on how to identify and exclude such distractions. We anticipate that by presenting ChatGPT-CARE with the most difficult example in the few-shot learning, the model will demonstrate competence in handling all cases effectively.

According to recent research, ChatGPT’s ability to provide detailed reasoning in response to clinical queries is limited^14,15^. Nonetheless, reasoning is critical in gaining trust of clinicians and assisting them in making decisions at the point of care. To address this, ChatGPT-CARE leveraged the chain-of-thought prompting technique to improve its reasoning capabilities. Prompt engineering refers to the systematic creation and optimization of prompts or instructions to influence the output of LLMs. Chain-of-thought prompting is a prompt engineering technique that involves a series of intermediate reasoning steps in prompts that simulate human thought processes in order to improve LLMs’ understanding and reasoning abilities. In ChatGPT-CARE, chain-of-thought prompting consists of two components: 1) checkpoint inference based on the input patient description; and 2) treatment path identification based on the Python-coded graph. In the first component, a patient description is provided, followed by a step-by-step example demonstrating how to infer responses for checkpoints (i.e., nodes in the graph) from the given patient description and medical knowledge. In the second component, the prompt demonstrates how to use these inferred nodes as inputs in the Python function in the system prompt to identify a path leading to the treatment option. Each of the 5-shot examples were represented following the chain-of-thought prompting strategy.

### Evaluation

To evaluate the effectiveness of ChatGPT-CARE in COVID-19 outpatient treatment decision support, we developed three distinct categories that describe various patient conditions, namely easy, medium, and hard. Each category comprised 15 descriptions, resulting in a total of 45 descriptions for evaluation. In the easy category, patient descriptions adhered closely to the exact phrases used in the COVID-19 Outpatient Treatment Guidelines. This allowed us to examine the capabilities of ChatGPT-CARE in making correct decisions at each checkpoint and identifying the correct treatment path in accordance with the Guidelines.

The medium category contained patient descriptions that used synonyms and semantically equivalent phrases to those in the Guidelines. For example, instead of describing a patient “who had a positive COVID-19 test”, the description was modified to “a positive Polymerase Chain Reaction (PCR) test”. The goal was to evaluate ChatGPT-CARE’s ability to comprehend the semantic meaning in the patient description, correctly map it to the relevant checkpoint, and make the correct decision.

The hard category included patient descriptions that differed considerably from the phrases used in the Guidelines and also contained information that required some level of inference. We also intentionally included unrelated and subtle information. The ChatGPT-CARE tool needs to not only extract symptoms from the description, but it also filters out irrelevant information. Through the hard category, we want to evaluate the capabilities of ChatGPT-CARE in inference, filtering out irrelevant information, and reasoning to make accurate treatment decisions. Examples for each category are shown in Table 1. We also note that the first 13 descriptions corresponded to different treatment paths in the Guidelines. The last two patient descriptions in each category were open-ended, designed by the physicians, and did not explicitly lead to a specific treatment path. These descriptions were used to assess ChatGPT-CARE’s generalizability.

**Table 1.** Example patient descriptions from easy, medium and hard categories.

| Patient Description: Easy | Patient Description: Medium | Patient Description: Hard |
| --- | --- | --- |
| *We have a patient who has a positive result for COVID-19, female, not pregnant, 17 years old and weighs 40 kg. The patient is at high risk for COVID-19 disease progression. She has a GFR of 36 mL/min. The patient is taking a drug that interacts with Paxlovid. Patient does not have authorization for outpatient infusion.* | *We have a 31-year-old thin patient testing positive for COVID-19. She weighs 40 kg. Patient has a history of genetic blood disorder. The patient has chronic kidney disease with eGFR=32 ml/min. She does not take other medications. And the patient has insurance's permission for remdesivir infusion.* | *We are presented with a patient who has tested positive with NAAT. The patient is a 19-year-old female weighing 42 kg and is not currently pregnant. It is noteworthy that the patient has undergone a surgical procedure involving the transplantation of healthy bone marrow stem cells to replace diseased or damaged bone marrow. The patient's glomerular filtration rate is measured at 29 ml/min. Moreover, the patient has not yet been granted authorization for the use of remdesivir.* |

Two physicians specialized in pulmonary disease and critical care evaluated the responses generated by ChatGPT-CARE on three aspects: accuracy, hallucination, and clarity. Accuracy assesses the correctness of the treatment decision result and the reasoning process that led to it. An accurate response provides the correct treatment for the patient. Hallucination refers to misinformation, unreasonable or illogical in common-sense knowledge^16^. In the responses generated by AI, hallucination can manifest as a fabricated premise or as unwarranted confidence in an obvious logical or mathematical error. Clarity evaluates the presence of ambiguity in responses, particularly in relation to entities or information referenced^17^. Table 2 lists available options for each aspect. The goal of the evaluation was to assess the effectiveness and reliability of ChatGPT-CARE responses in a clinical setting. The agreement between the two physicians was calculated based on the percentage of overlap.

**Table 2.** Three evaluation aspects and available values and descriptions.

| Evaluation Aspects | Values | Description |
| --- | --- | --- |
| Accuracy | Accurate | The treatment recommendation and decision-making process are correct. |
|  | Acceptable | The response is deemed helpful but not entirely correct. |
|  | Inaccurate | The response is considered misleading and unhelpful. |
| Hallucination | Present | Hallucination is present in the response. |
|  | Absent | Hallucination is absent in the response. |
| Clarity | Comprehensive | The response is deemed reasonable, detailed, and free from ambiguity. |
|  | Understandable | The response is deemed reasonable but not exhaustive. |
|  | Vague | The response is ambiguous, irrelevant or unclear. |

### Model Selections and Parameters

We implemented both GPT-3.5 (gpt-3.5-turbo-16k) and GPT-4 through OpenAI API in ChatGPT-CARE and compared the responses with the two versions of ChatGPT, denoted as ChatGPT-CARE (GPT-3.5), ChatGPT-CARE (GPT-4), ChatGPT (GPT-3.5), and ChatGPT (GPT-4), respectively. We set the temperature to 1 to maximize creativity in the responses and maintain the other parameters at their default values.

## Results

The overall agreement between the two physicians is 64%, specifically, 70% for accuracy, 74% for hallucination, 49% for clarity, which indicates moderate agreement. The physicians’ evaluations of accuracy, hallucinations, and clarity of responses from the two ChatGPT-CARE models and the two ChatGPT models are depicted by stacked bar charts in Figure 2. Responses from ChatGPT-CARE (GPT-3.5) and ChatGPT-CARE (GPT-4) were judged to be more accurate and clearer than ChatGPT (GPT-3.5) and ChatGPT (GPT-4) respectively for all three patient categories. In particular, ChatGPT-CARE (GPT-4) generated the most accurate responses for all three categories. On the other hand, ChatGPT (GPT-3.5) and ChatGPT (GPT-4) generated responses with poor clarity and were less accurate in the hard and medium categories. ChatGPT-CARE (GPT-4) responses were judged to be more comprehensive than ChatGPT-CARE (GPT-3.5) responses for hard and medium categories, while responses for the easy category are slightly less comprehensive. This finding is consistent with previous studies^3^ suggesting that GPT-4 possesses better inference capabilities compared to GPT-3.5.

**Figure 2.**
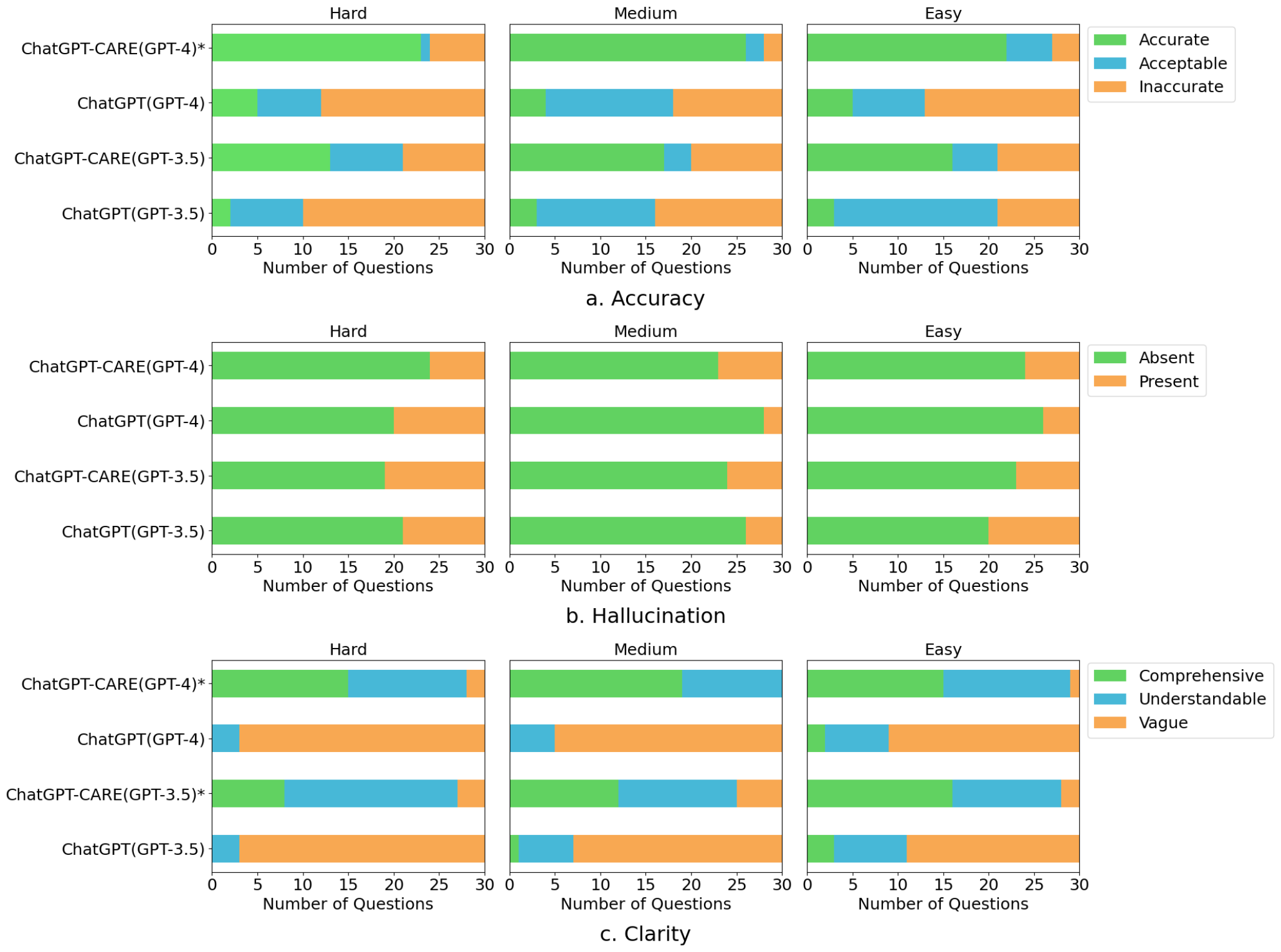
Physician evaluations of ChatGPT-CARE in accuracy, hallucination and clarity. * indicate the difference between ChatGPT-CARE and original ChatGPT is statistically significant (p < 0.01).

The findings on hallucination are mixed. Both versions of ChatGPT-CARE demonstrated reduced hallucination compared to the respective versions of ChatGPT. However, ChatGPT-CARE (GPT-4) tended to produce more hallucinations for medium and easy categories compared to ChatGPT (GPT-3.5). On further review, we discovered that the ChatGPT versions, while unable to provide completely satisfactory responses for medium and easy descriptions, prevented hallucinations by providing brief and vague responses. In contrast, both versions of ChatGPT-CARE deduce responses from specific details in the patient descriptions, which occasionally results in hallucinations.

Nonetheless, it is important to note that the hallucination rate remains low across all four models. We identified two major categories of hallucinations: medical knowledge hallucinations and mathematical hallucinations. All four models produce hallucinations in the medical knowledge, such as “genetic blood disorder” falling under “sickle cell disease” (sickle cell disease is a genetic blood disorder that falls under the category of hemoglobinopathies) and “Remdesivir is absolutely contraindicated in patients with end-stage renal disease” (Remdesivir is not absolutely contraindicated). Most mathematical hallucinations are produced by ChatGPT (GPT-3.5) and ChatGPT-CARE (GPT-3.5), such as the classification of a patient weighing more than 40 kg as weighing less than 40 kg. The GPT-4 based models have significantly fewer mathematical hallucinations.

We present examples generated by ChatGPT-CARE (GPT-4) to demonstrate its inference and reasoning capabilities, as depicted in Figure 3. For easy category descriptions that adhere closely to the Guidelines, the model effectively identifies nodes and makes decisions without difficulty. The medium category example used synonyms like “asymptomatic”, so the model determined that the patient did not have symptoms and assigned them to low risk. In the hard category example, the model demonstrated two key inferences. First, it deduced from the phrase “testing positive for NAAT” that NAAT refers to a type of COVID test, concluding that the patient had a positive COVID test. Second, it determined that an estimated glomerular filtration rate (eGFR) of 32 ml/min falls under the category “>30 ml/min,” leading the model to the node “Patient doesn’t have severe renal impairment (GFR>30 ml/min). These inference and reasoning capabilities are very powerful, which typically require multiple techniques, such as information extraction and classification, and several steps in conventional NLP.

**Figure 3.**
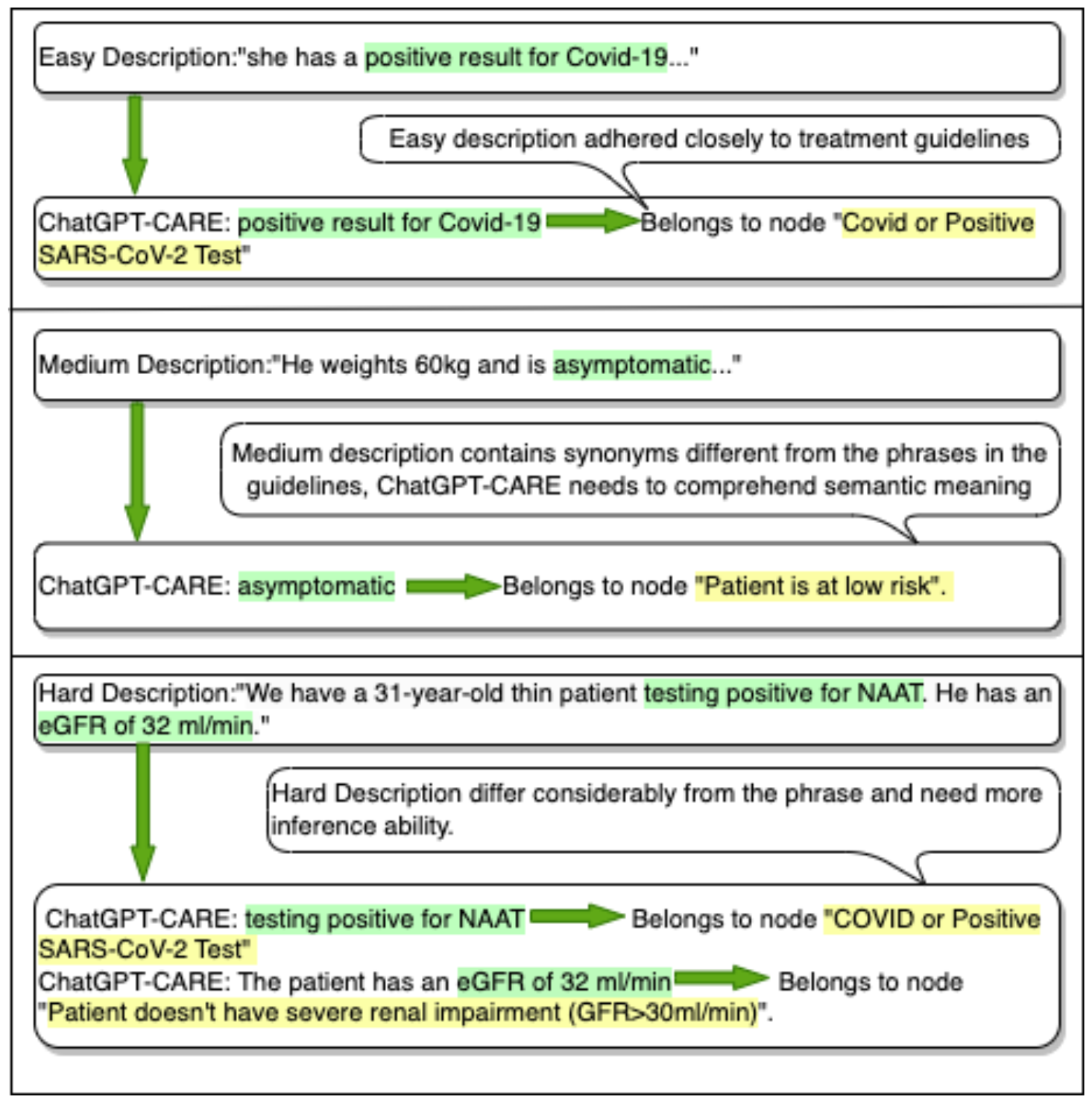
Inference and reasoning capabilities of ChatGPT-CARE. Phrases from the input patient descriptions are highlighted in green; inferences made by ChatGPT-CARE are highlighted in yellow.

## Discussion

In this study, we introduce ChatGPT-CARE, a tool that integrates clinical practice guidelines with ChatGPT to enhance clinical decision support. Although the tool focuses on outpatient treatment decisions for COVID-19, the proposed method could be easily applied to other clinical decision-making processes. ChatGPT-CARE utilizes in-context learning to fine-tune the model for COVID-19 treatment decision support. Chain-of-thought prompting is employed to improve ChatGPT-CARE’s reasoning capabilities. Evaluation of the tool includes three categories of patient descriptions representing different levels of complexity in patient conditions and two physicians were asked to compare and evaluate the responses generated by ChatGPT-CARE and the original ChatGPT for accuracy, hallucination, and clarity. The results demonstrate ChatGPT-CARE’s higher accuracy and clarity compared to ChatGPT in all patient description categories, showcasing its improved inference capabilities. Overall, ChatGPT-CARE outperforms other models in generating accurate treatment recommendations, leveraging its advanced inference, and reasoning capabilities. Utilizing its advanced inference and reasoning capabilities, ChatGPT-CARE outperforms other models in generating accurate treatment recommendations.

In ChatGPT-CARE, we defined a Python function to determine the treatment path based on given nodes. This Python function was executed in real-time within ChatGPT-CARE without the need for a separate Python interpreter. Initially, we tested the function externally using a Python interpreter outside ChatGPT-CARE. However, during subsequent tests, we discovered that some descriptions yielded null results when executed outside ChatGPT-CARE. The root cause of this issue was traced back to our Python function’s requirement for a list of nodes as input, which should not include nodes from two different paths in the graph. However, ChatGPT-CARE sometimes incorrectly inferred nodes from the description that may result in two paths. Interestingly, when the Python function was executed inside ChatGPT-CARE, it managed to return correct results in most cases, even when some nodes came from different paths. This observation suggests that ChatGPT-CARE exhibits a certain level of flexibility in handling mixed nodes, prioritizing the majority of nodes on the correct path to achieve accurate outcomes. However, this success within ChatGPT-CARE comes at the cost of potential inconsistencies because it does not adhere strictly to the mechanics of a Python interpreter. As a result, there might be instances where ChatGPT-CARE’s behavior differs from what one would expect from a traditional Python environment. Additional research is needed to further investigate this phenomenon and its underlying reasons.

There are 2 open-ended patient descriptions in each category. They were proposed by our physicians to evaluate the model’s adaptability to incomplete and uncommon information without predetermined correct answers. In response to these open-ended questions, it is consistent with the previous conclusion that ChatGPT-CARE outperforms the original ChatGPT in terms of accuracy and clarity. However, it is important to note that when faced with incomplete descriptions lacking crucial details of making decisions for COVID-19 treatment, both models make arbitrary assumptions regarding those missing elements. For instance, there is an open-ended description that do not mention the patient’s access to Redemsivir, and ChatGPT-CARE (GPT-3.5) assumes the availability of Redemsivir to the patient, while ChatGPT-CARE (GPT-4) assumes its unavailability. To evolve into a robust tool, ChatGPT-CARE needs to proactively prompt the user to provide key information or offer varied recommendations based on distinct situations, rather than making arbitrary assumptions.

We also interviewed the two physicians who evaluated the ChatGPT-CARE and ChatGPT. Figure 4 shows their feedback on these models. The physicians were notably impressed with ChatGPT-CARE’s accuracy, specificity, and clarity in providing responses. They like that the ChatGPT-CARE models can provide valuable information in determining treatment options, particularly for complex patients. On the other hand, the original ChatGPT primarily delivered general responses, occasionally containing inaccuracies and lacking specificity. They believed the original ChatGPT could be useful for the general public but was insufficient for assisting physicians in making crucial decisions.

**Figure 4.**
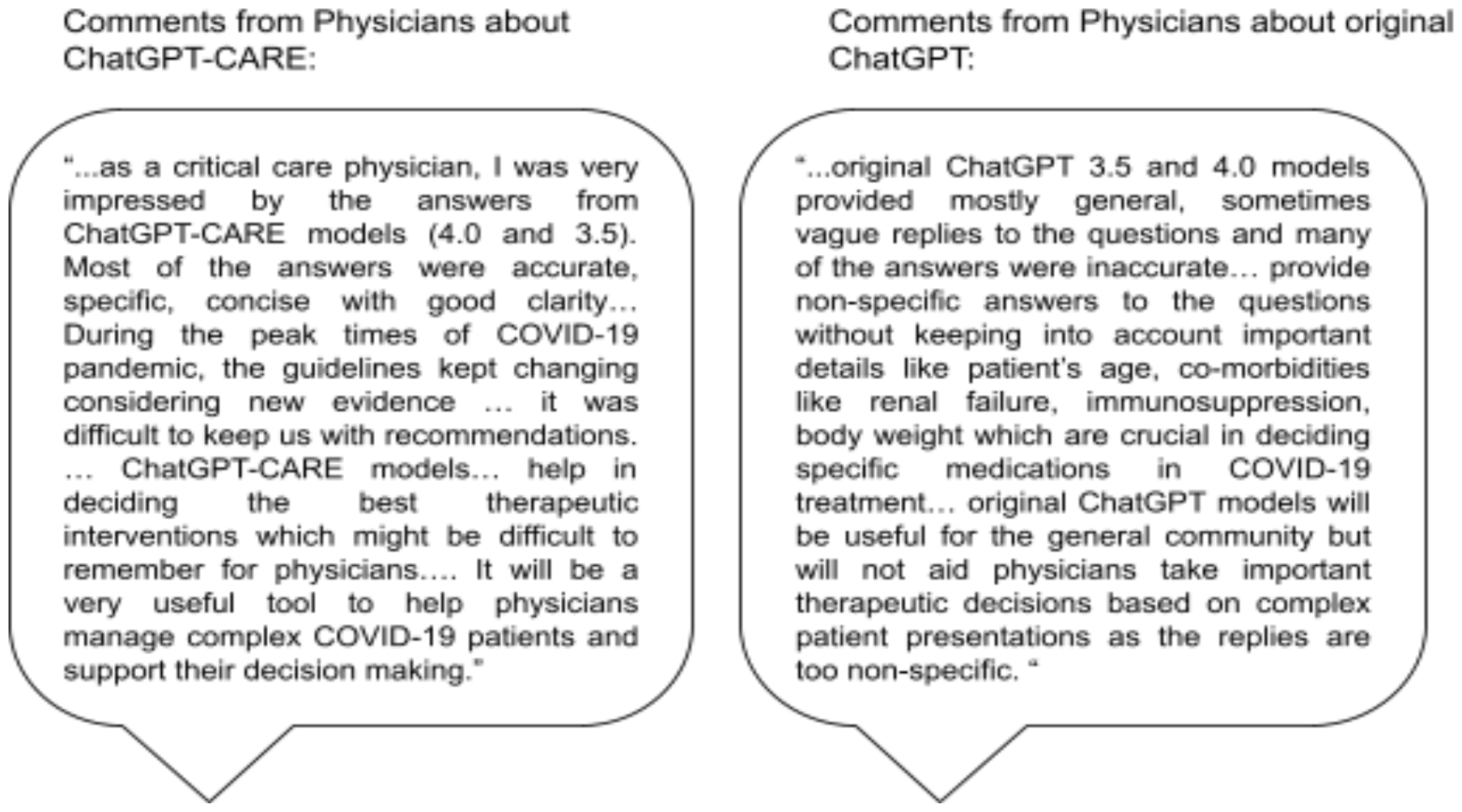
Interview comments from physicians about ChatGPT-CARE and ChatGPT.

In clinical settings, where decisions can have life-or-death consequences, it is crucial to be able to trust and comprehend the reasoning behind an AI tool’s recommendation. Transparency ensures that clinicians can follow the model’s logic and evidence, aligning it with clinical practice guidelines and ethical concerns. However, current limitations in deep learning-based decision support tools include a lack of transparency and nuanced understanding of complex medical scenarios, as well as difficulties integrating the most recent clinical practice guidelines. Explaining the reasoning in human-understandable terms, such as in ChatGPT-CARE, are essential steps in overcoming these limitations and making LLMs a valuable tool in clinical practice.

We are developing ChatGPT-CARE as a ChatGPT plugin in order to make this tool widely accessible. Recognizing the unique demands and sensitivities of the healthcare environment, the ChatGPT-CARE plugin will be designed not only with a focus on accuracy and reasoning but also with strong ethical considerations, guided by the GREAT PLEA principles^18^ that our team has developed. We are committed to rigorous testing and validation to ensure that it meets the highest standards of quality and reliability. We believe that ChatGPT-CARE will mark a significant advancement in the integration of generative AI technologies in clinical practice.

## Data Availability

All data produced in the present study are available upon reasonable request to the authors

